# Impact of COVID-19 Diagnosis on Weight Trajectories of Children in the US National COVID Cohort Collaborative

**DOI:** 10.1101/2025.01.03.25319927

**Authors:** Md Mozaharul Mottalib, Thao-Ly T. Phan, Carolyn T Bramante, Christopher G Chute, Lee A Pyles, Rahmatollah Beheshti, N3C

## Abstract

**Background:** The COVID-19 pandemic has exacerbated the obesity epidemic, with both adults and children demonstrating rapid weight gain during the pandemic. However, the impact of having a COVID-19 diagnosis on this trend is not known.

**Methods:** Using longitudinal data from January 2019 to June 2023 collected by the US National Institute for Health’s National COVID Cohort Collaborative (N3C), children (age 2–18 years) with positive COVID-19 test results (n=11,474, 53% male, mean [SD] age 5.57 [±3.29] years, 54% white, mean [SD] 5.2 [±2.9] BMI observations per participants) were matched with COVID-19 negative children with identical demographic characteristics and similar observation window. We compared BMI percentile trajectories between the COVID-19 positive and COVID-19 negative cohorts, with further evaluation performed on COVID-19 positive patients stratified by hospitalization status.

**Results:** COVID-19 positive patients had a greater increase in %BMI_p95_ than COVID-19 negative patients (average increase of 2.34 (±7.73) compared to 1.46 (±6.09), p<0.0005). COVID-19 positive patients gained more weight after their diagnosis of COVID-19 than before. Non-hospitalized children gained more weight than hospitalized children (average increase in %BMI_p95_ of 2.38 (±7.65)) compared to 1.87 (±8.54)). Mixed effect regression analyses demonstrated that these associations remained even after adjusting for time, demographics, and baseline %BMI_p95_.

**Conclusions:** Having a COVID-19 diagnosis was associated with more rapid weight gain, especially after diagnosis and early in the pandemic. Future research should explore the reasons for this association and the implications for future health emergencies.

**Authors’ Contributions:** M.M.M. drafted the initial manuscript and assisted with the data curation, formal analysis, methodology, and visualization. T.T.P. assisted with the study design, interpretation of results, funding acquisition, and manuscript drafting and editing. C.T.B. and C.G.C assisted with formal analysis and interpretation of results. L.A.P. assisted with the interpretation of results and manuscript editing. R.B. supervised the study with study design, resources, funding acquisition, and manuscript editing. All authors assisted in the conceptualization of the study and approved it for publication.

**Author Disclosure Statement:** The authors declare no conflicts of interest.

**Impact Statement:** This study investigating the weight trajectories of children during the COVID-19 pandemic shows that the pandemic has led to a significant increase in weight gain among US children aged 6-11, with boys gaining more weight than girls. Non-hospitalized COVID-19-positive children were found to be at greater risk of gaining weight.

**Declarations:** The analyses described in this publication were conducted with data or tools accessed through the NCATS N3C Data Enclave https://covid.cd2h.org and N3C Attribution & Publication Policy v 1.2-2020-08-25b supported by NCATS U24 TR002306, Axle Informatics Subcontract: NCATS-P00438-B, [DUR RP-0BDD0E] and was supported by NIH awards, P20GM103446 and U54-GM104941. This research was possible because of the patients whose information is included within the data and the organizations (https://ncats.nih.gov/n3c/resources/data-contribution/data-transfer-agreement-signatories) and scientists who have contributed to the ongoing development of this community resource [https://doi.org/10.1093/jamia/ocaa196]

## Introduction

The global impact of the severe acute respiratory syndrome coronavirus 2 (SARS-CoV-2), later designated as Coronavirus Disease 2019 (COVID-19), pandemic has been profound. To date, it has afflicted over 750 million individuals, resulting in a staggering toll of more than 7 million lives lost(1, 2). While its impact on adults has been extensively studied(3–5), there has been limited exploration of its effects on children and adolescents(6–8). However, the pandemic-induced shifts in lifestyle behaviors could have enduring effects on children’s weight(9). As a response to the pandemic, public safety measures such as lockdowns, quarantine, and social distancing recommendations disrupted the daily lives of families and children. These unique challenges forced families to adapt to new circumstances, prompting changes in their lifestyles and eating behaviors. Increased sedentary behavior, extended screen time, and reduced physical activity have been associated with a rise in obesity across populations(10–14). Not long after the start of the pandemic, the American Academy of Pediatrics (AAP) raised concerns that these changes may adversely impact children’s nutrition, physical activity, and obesity risk(15) and exacerbate an existing obesity epidemic that affects nearly 20% of children in the United States(16, 17) and poses substantial risks for comorbid conditions, including cardiometabolic diseases and mental health problems(18–21).

Studies have consistently demonstrated significant increases in weight among children and adults during the pandemic. For example, Lin et al.(3) conducted a longitudinal cohort study of adults during the initial shelter-in-place order in the US from March 19, 2020, to April 6, 2020. They found a significant increase in weight over a 3-month period at a rate of about 1.5 lbs. weight gain per month. Woolford et al.(22) compared the BMI of youth aged 5 to 17 years during the pandemic in 2020 with the BMI in the same period before the pandemic in 2019 and found significant increases in rates of overweight and obesity in 2020(4, 23–25). Since then, several pediatric studies have elucidated potential lifestyle, behavioral, and psychosocial causes of weight change among children due to the pandemic(26–29) and have confirmed significant increases in weight during the pandemic that exceeded the usual pre-pandemic weight change(6, 22, 30–32).

While it is clear that the pandemic has led to significant weight gain among children, studies have not examined the impact of having a COVID-19 diagnosis on this trend. Indeed, research related to COVID-19 diagnoses has been limited by the paucity of large, multi-institutional datasets of individuals testing positive for COVID-19(33–36). In this study, we leverage the extensive database of the National COVID Cohort Collaborative (N3C),(37) which includes patients from across the country who tested positive for COVID-19 and demographically matched controls. Our study presents an in-depth analysis of the changes in children’s weight during this unprecedented time, especially focusing on understanding how a COVID-19 infection and hospitalization for a COVID-19 infection impacted weight trajectories among children during different stages of the pandemic(38).

## Materials and Methods

### National COVID Cohort Collaborative (N3C)

National COVID Cohort Collaborative (N3C) is a multisite partnership that is comprised of members from the NIH Clinical and Translational Science Awards Program, it’s Center for Data to Health, the IDeA Centers for Translational Research, and several Electronic Health Record (EHR)-based research networks.(39) It aggregates and harmonizes electronic health record (EHR) data across clinical organizations and health system entities in the US to create a longitudinal multicenter cohort of patients with a COVID-19 diagnosis, matched in a 2:1 ratio to patients without a COVID-19 diagnosis by site, age, sex, and race. N3C has a national governance and regulatory body, uses COVID-19 cohort definitions via community-developed phenotypes, harmonizes data across 4 common data models (CDMs), and has a collaborative analytics platform to support the deployment of novel algorithms of data aggregated from the United States(37). The N3C has unique features that distinguish it from other COVID-19 data resources. It harmonizes data from many clinical sites across the US (98 signed data transfer agreements as of February 2024). This is important since many US reports of COVID-19 clinical characteristics come from a single hospital or healthcare system in a single geographic region. Furthermore, central curation ensures that N3C data are robust, and the quality is assured across sites. The N3C database contains data on weight, height, BMI, BMI percentile, and obesity diagnosis. These codes are adjusted and synchronized by the N3C regulatory team for easier analysis(40).

### Data Extraction and Cohort Definition

We accessed the level 3 Limited Data Set (LDS) from the N3C data enclave, which includes patient data with dates of service and patient ZIP code. This data is only available to US-based research institutes with approved Data Use Request (DUR). We performed a retrospective analysis of children undergoing COVID-19 testing at an N3C site between January 1, 2019 (start date of N3C database) and June 30, 2023 (date of data extraction), whose data were completely harmonized, integrated, and released for analysis. Patients were included as COVID-19 positive cases if they met the following criteria: 1) aged 2 to 18 years old, 2) tested positive for COVID-19 following the N3C phenotyping guidelines to define COVID-19 positivity based on a COVID-19–positive polymerase chain reaction or antigen test, or an International Classification of Diseases (ICD)-10-CM diagnostic code for COVID-19 during the same single index encounter(41), and 3) had at least two BMI measurements or body height/weight measurements, with at least one measurement within 6 months before diagnosis and one measurement within 6 months after diagnosis. From an initial population of 521,903 patients (adult and pediatric), 25,936 patients were found to meet the criteria (**Figure 1**).

**Figure 1.**
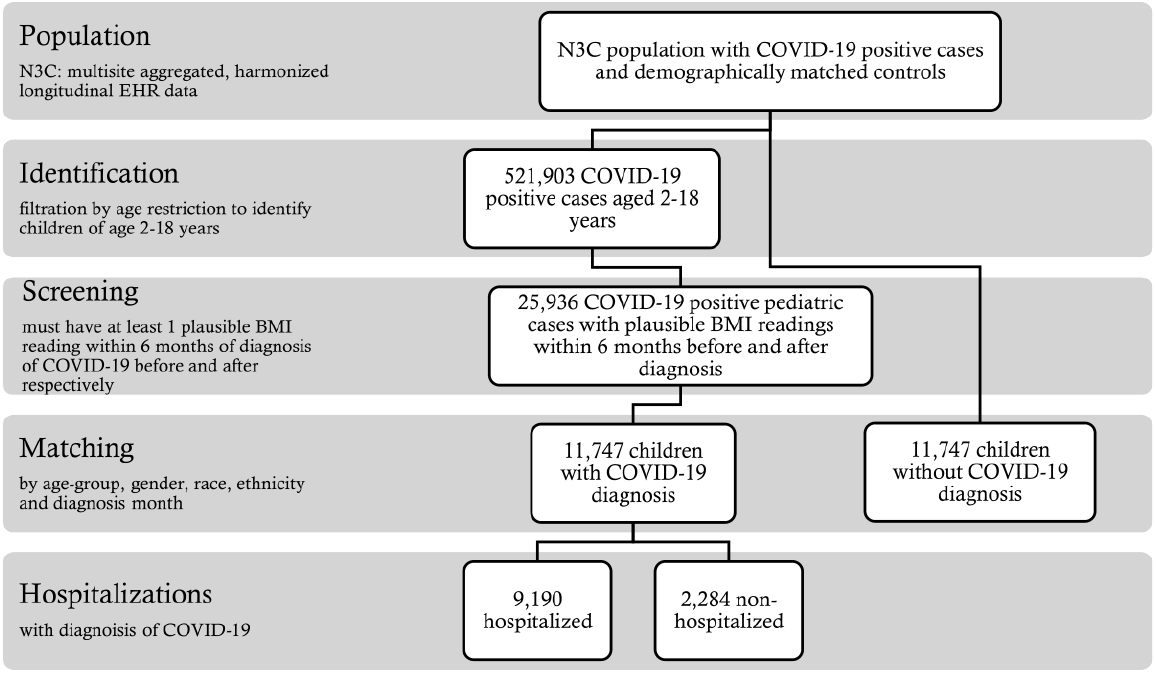
Flowchart of cohort selection.

We then matched the cohort with patients without a diagnosis of COVID-19 in the repository (controls, COVID-19 negative patients), based on age group, gender, race, ethnicity, and observation window. For example, a positive-diagnosis patient with observations from November 2019 to November 2020 was matched with a negative-diagnosis patient having observations starting from November 2019, spreading over a 13-month observation window. This reduced each cohort to 11,474, as not every matched control had two or more weights and heights in the database within a 13-month timeframe. Matching was done using the R package MatchIt(42) with a generalized linear model for calculating the propensity score among the samples. Matching was performed with a greedy nearest neighbor matching algorithm implemented in the MatchIt package. The choice of this algorithm yielded the maximum number of matches, keeping the cohort considerably large. In the initial population, only around 32% of the patients had more than 2 measurements. However, in the final matched cohort, there were around 46% of the patients who had more than 2 measurements.

### Demographic Characteristics

For the cohort defined above, demographic characteristics were extracted, including date of birth (to derive age), sex, race, and ethnicity. For the purposes of analysis, age, sex, race, and ethnicity were grouped into broad categories (**Table 1**). It is important to note that the COVID-19 negative (control) group exhibited identical demographic characteristics and had weight measurements available at comparable time points for each corresponding patient.

**Table 1.** Average change in %*BMI*_*p*95_ before and after the COVID-19 pandemic periods in patients both with and without COVID-19 diagnosis aged 2-18 years with measurements available for about a year. [Early pandemic: January 2020 – December 2020, Late pandemic: January 2021 – June 2023, (*) represents statistically significant difference with a p-value < 0.005]

| Characteristics | Average change in %BMI<sub>p95</sub> (±SD) |  |  |  |  |  |
| --- | --- | --- | --- | --- | --- | --- |
|  | Control |  |  | Covid positive |  |  |
|  | Early pandemic | Late pandemic | Difference | Early pandemic | Late pandemic | Difference |
| **Overall** | 1.23 (±0.15) | 1.87 (±0.27) | 0.64* | 2.44 (±0.29) | 2.21 (±0.2) | - 0.23* |
| **Age group** |  |  |  |  |  |  |
| 2-5 (60.14%) | 1.45 (±0.23) | 2.26 (±0.40) | - 0.81* | 1.82 (±0.39) | 1.69 (±0.26) | - 0.13* |
| 6-11 (33.29%) | 0.96 (±0.18) | 1.23 (±0.33) | - 0.27 | 3.44 (±0.49) | 3.32 (±0.34) | - 0.12* |
| 12-15 (4.75%) | 1.01 (±0.52) | 1.30 (±1.28) | - 0.29 | 3.08 (±1.00) | 2.06 (±1.08) | - 1.02* |
| 16-18 (1.82%) | - 0.13 (±1.10) | 1.52 (±1.93) | 1.65 | 0.945 (±2.50) | -0.64 (±1.83) | - 1.58 |
| **Gender** |  |  |  |  |  |  |
| Male (53.28%) | 1.27 (±0.21) | 1.63 (±0.36) | 0.36* | 3.19 (±0.40) | 2.65 (±0.27) | - 0.57* |
| Female (46.71%) | 1.19 (±0.22) | 2.14 (±0.42) | 0.95* | 1.56 (±0.42) | 1.73 (±0.29) | 0.17* |
| **Race** |  |  |  |  |  |  |
| White (53.41%) | 1.11 (±0.18) | 1.88 (±0.33) | 0.77* | 2.72 (±0.56) | 2.09 (±0.43) | - 0.02* |
| Black or African American (17.93%) | 1.82 (±0.37) | 1.78 (±0.57) | - 0.03* | 3.19 (±0.62) | 2.37 (±0.45) | - 0.82* |
| Asian (3.33%) | 0.70 (±0.65) | 0.89 (±1.60) | 0.19 | 1.99 (±1.61) | 1.88 (±1.07) | - 0.12 |
| Native Hawaiian or Other Pacific Islander (0.1%) | 2.33 (±4.28) | - 0.29 (±4.05) | - 4.35 | 1.49 (±5.29) | 1.99 (±2.53) | 0.49 |
| Other (25.23%) | 0.94 (±0.53) | 2.48 (±1.48) | 1.54 | 2.36 (±0.95) | 1.76 (±0.68) | - 0.6 |
| **Ethnicity** |  |  |  |  |  |  |
| Hispanic or Latino (28.66%) | 1.27 (±0.32) | 1.86 (±0.59) | 0.59* | 2.72 (±0.56) | 2.09 (±0.43) | - 0.63* |
| Non-Hispanic or Latino (66.21%) | 1.22 (±0.17) | 1.87 (±0.31) | 0.65* | 2.33 (±0.34) | 2.24 (±0.23) | - 0.09* |

### Weight Status Metrics

For the cohort defined above, we extracted height and weight measurements starting from 12 months before the COVID-19 diagnosis up until the date of data extraction. Weights were restricted between 5kg and 300kg and heights between 0.6m to 2.43m according to CDC growth parameters established for the defined age range(43). BMI was calculated using, *BMI* = *weight/height*^2^ formula as *kilogram/meter*^2^. Extreme values of BMI were excluded based on their modified z-scores(44).

In the U.S., the CDC 2000 BMI charts are recommended for use in children 2 to 20 years of age to determine a child’s BMI percentile in relation to norms for age and sex. Freedman et al.(45) illustrated the challenges in using BMI percentiles in children of different ages and compared alternative metrics to quantify adiposity status for children with a BMI at the upper extremes. Because of these challenges, many in the research and clinical community use percent above the 95^th^ percentile for those with extreme BMI, with 120% of the 95th percentile of BMI (%*BMI*_*p*95_) is used to define severe obesity in children (46). The percent of the BMI 95^th^ percentile is defined as:>

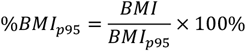

where, *BMI*_*p*95_ shows the BMI 95^th^ percentile. Because of the common limitations of EHR data (i.e., missingness and irregular sampling), *BMI*_*p*95_ values were needed to be imputed to derive trajectories. Only 2 measurements would represent a change score instead of a trajectory. Polynomial interpolation of order 2 was used to impute the missing values, where at least 4 measurements were present (∼40% of the cohort) in the trajectory; for the remainder of cases, linear imputation was used. In total around 10,500 patient trajectories (∼46% of the cohort) were imputed using the aforementioned techniques.

### Statistical Methodologies

Descriptive statistics were summarized by the mean and standard deviation (SD) for continuous variables and frequency with percent for categorical variables. One-way ANOVA was used for comparing continuous variables, and the chi-squared test was used for categorical variables, implemented using the Python Statsmodels(47) and SciPy(48) libraries. We generated figures depicting the weight trajectories of the COVID-19 positive and COVID-19 negative cohorts, stratified by demographic groups. We performed several statistical analyses on the mean %*BMI*_*p*95_ changes during the Early Pandemic (January 2020 – December 2020) and Late Pandemic (January 2021 – June 2023). Bartlett testing was performed to evaluate the homogeneity of variance among the mean %*BMI*_*p*95_ change(49). Since the variances were not equal among groups based on stages of the pandemic, Kruskal-Wallis H-test(50) was performed on the change in %*BMI*_*p*95_. We used mixed-effects models to examine the rates of change in the %*BMI*_*p*95_, to examine the impact of having a COVID-19 infection on change in %*BMI*_*p*95_. Additionally, a similar but separate examination of weight change in the COVID-19 positive patients was performed to examine the impact of having a hospitalization on change in %*BMI*_*p*95_. For both models, we included time, baseline %*BMI*_*p*95_ and demographics as covariates. The time of COVID-19 diagnosis, especially the pandemic era indicator, was an important variable in the analyses where we investigated the weight change trend during different pandemic eras. Demographics were included as fixed effects in the models.

## Results

### Comparison of BMI Percentile Trajectories between COVID-19 Positive and COVID-19 Negative Cohorts

%*BMI*_*p*95_ trajectories of COVID-19 positive cases and COVID-19 negative controls are illustrated in **Figure 2**, with stratification by demographic groups in **Figure 3**. Baseline %*BMI*_*p*95_ was higher in the COVID-19 positive group (mean 88.07, SD 7.25) compared to the COVID-19 negative group (mean 85.11, SD (10.31, p < 0.005) except among the youngest children (aged 2-5 years). Although the actual difference was 2.96 percentile points, even modest increases in BMI percentile among children, particularly those already in higher percentiles, are clinically meaningful in the context of obesity and associated health risks, such as more severe COVID-19 outcomes. Both groups demonstrated an increase in %*BMI*_*p*95_ over the first 6 months of the time period, but only COVID-19 positive patients continued to have an increase in %*BMI*_*p*95_ in the latter 6 months, after diagnosis. In fact, COVID-19 positive patients gained more weight after their diagnosis of COVID-19 than before the diagnosis (average increase in weight of 2.34 (±7.73) after diagnosis vs. 1.46 (±6.09) before diagnosis, p < 0.005). Children between the ages of 6 and 11 demonstrated more rapid increases in %*BMI*_*p*95_ than other age groups, especially among the COVID-19 positive cohort. Male patients in the COVID-19 positive cohort demonstrated more rapid increases in %*BMI*_*p*95_ than female patients. There were no major differences in BMI percentile trajectories by race or ethnicity. Mixed effects regression analyses showed that the association between having a COVID-19 diagnosis and having a more rapid increase in %*BMI*_*p*95_ remained, even when accounting for demographics, time, and baseline %*BMI*_*p*95_ (coefficient = 31.05, p= 0.005). In the regression model, only age group and gender also had a significant association with change in %*BMI*_*p*95_.

**Figure 2.**
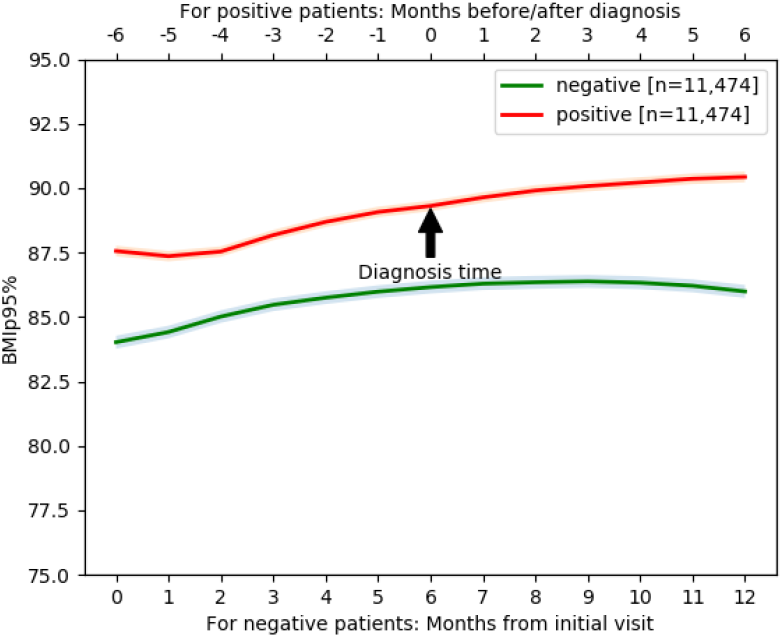
%*BMI*_*p*95_ trajectory with 95% confidence interval band over 13-month period including 6-months prior to diagnosis and 6-month after diagnosis for COVID-19 positive cases and over the same 13-month period for matched COVID-19 negative patients.

**Figure 3.**
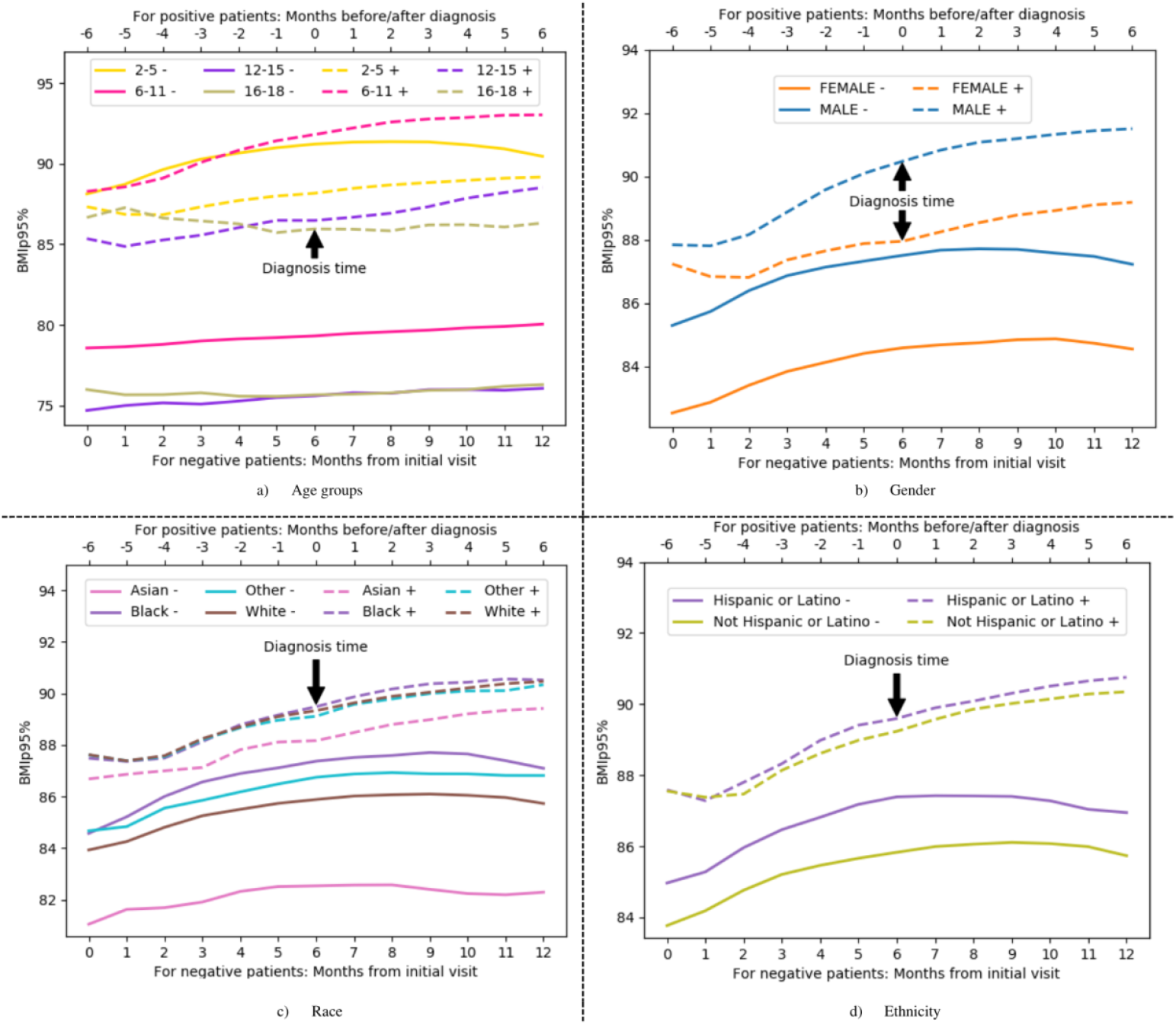
%*BMI*_*p*95_ trajectories based on demographic group. Each line represents the mean value at that time period.

### Comparison of Change in BMI Percentile between COVID-19 Positive and COVID-19 Negative Cohorts Based on Pandemic Stage

**Table 1** presents the average change in %*BMI*_*p*95_ for the COVID-19 positive and negative cohorts by stages of the pandemic. Both cohorts had an increase in %*BMI*_*p*95_ during the early and late pandemic stages, but patients in the COVID-19 positive cohort had greater increases in both early (average %BMI_p95_ increase of 2.44 percentiles) and late pandemic (average %*BMI*_*p*95_ increase of 2.21 percentiles) stages compared to the COVID-19 negative cohort (average %*BMI*_*p*95_ increase of 1.23 and 1.87, respectively). The COVID-positive cohort gained more weight during the early pandemic compared to the late pandemic, whereas the COVID-negative cohort gained more weight during the late pandemic compared to the early pandemic. These trends were the same for patients regardless of demographic group, except for Black patients without a COVID-19 diagnosis and female patients with a COVID-19 diagnosis.

### Comparison of Change in BMI Percentile between COVID-19 Positive Patients Based on Hospitalization

Approximately 19% of children among the COVID-19 positive cohort were hospitalized. As shown in **Figure 4**, non-hospitalized children had greater changes in BMI percentile over time (average %*BMI*_*p*95_ change of 2.38 (±7.65)) compared to hospitalized children (average increase of 1.87 (±8.54) %*BMI*_*p*95_). Hospitalized children were more likely to be younger and Black (**Table 2**). Mixed effects regression analyses showed that the association between being hospitalized for COVID-19 and less rapid increase in %*BMI*_*p*95_ remained, even when accounting for demographics, time, and baseline %*BMI*_*p*95_ (coefficient = 29.684, 95% p = 0.003). In the regression model, only age group and gender also had a significant association with change in %*BMI*_*p*95_.

**Figure 4.**
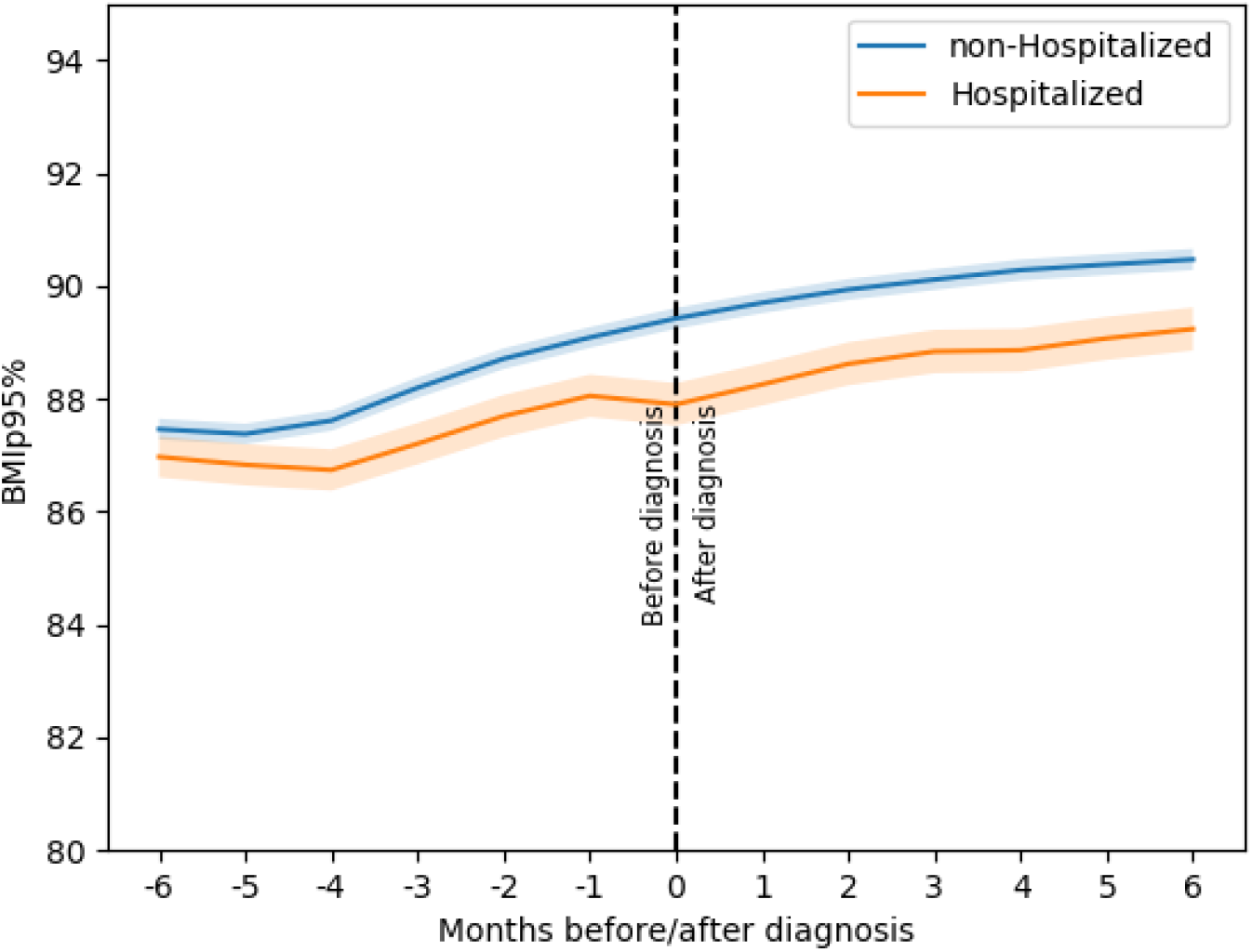
%*BMI*_*p*95_ trajectories based on hospitalization. Each line representing the trajectory of the mean values at specific time periods, the shaded region represents the 95% confidence interval.

**Table 2.** Demographic characteristics of cohort based on hospitalization status.

| Characteristics | Patients, no. (%) |  | Hospitalization Risk Ratio | p-value |
| --- | --- | --- | --- | --- |
|  | Not hospitalized (n = 9190) | Hospitalized (n = 2284) |  |  |
| Age |  |  |  | < 0.005 |
| Age (y), mean (SD) | 5.63 (3.287) | 5.24 (3.22) | - |  |
| 2-5 | 5433 (59.12%) | 1443 (63.18%) | Ref. |  |
| 6-11 | 3134 (34.1%) | 705 (30.87%) | 0.88 |  |
| 12-15 | 455 (4.95%) | 111 (4.86%) | 0.93 |  |
| 16-18 | 168 (1.83%) | 25 (1.09%) | 0.62 |  |
| Gender |  |  |  | 0.0402 |
| Male | 4843 (52.7%) | 1270 (55.6%) | Ref. |  |
| Female | 4346 (47.29%) | 1014 (44.4%) | 0.91 |  |
| Race |  |  |  | < 0.005 |
| White | 5004 (54.45%) | 1125 (49.26%) | Ref. |  |
| Black or African American | 1567 (17.05%) | 491 (21.5%) | 1.30 |  |
| Asian | 286 (3.11%) | 59 (2.58%) | 0.93 |  |
| Other | 666 (7.25%) | 174 (7.62%) | 1.13 |  |
| Ethnicity |  |  |  | 0.0207 |
| Hispanic or Latino | 2607 (28.37%) | 702 (30.74%) | Ref. |  |
| Non-Hispanic or Latino | 6109 (66.47%) | 1487 (65.11%) | 0.92 |  |

## Discussion

While much has been written about the impact of the COVID-19 pandemic on weight gain among children in the United States, this is the first study that we are aware of that highlights how having a diagnosis of COVID-19 contributed to this trend. Using a large nationally representative database, we found that children with a diagnosis of COVID-19 had larger and more persistent increases in BMI percentile over time compared to children without a COVID-19 diagnosis, especially early in the pandemic. Children with a diagnosis of COVID-19 gained more weight after diagnosis than before diagnosis. We also found that among children with a COVID-19 diagnosis, those who were hospitalized had less rapid weight gain than those who were not hospitalized. These findings add to our understanding of the weight gain experienced by children during the pandemic and highlight the complexity of factors that likely contributed.

Our study leverages the National COVID Cohort Collaborative (N3C), which includes nearly 100 institutions working together to build a large, centralized data resource to study COVID-19. Under the stewardship of the NIH’s National Center for Advancing Translational Science, N3C has harmonized EHR data across health systems and uniquely consists of cases (patients with a COVID-19 diagnosis) and demographically matched controls without a COVID-19 diagnosis. Because of this, we can uniquely comment on a nationally representative sample and on the impact of a COVID-19 diagnosis and hospitalization for COVID-19 on weight outcomes. To our knowledge, there exist only a few studies in adults that have examined this question and no studies in pediatric populations.

As noted in the Introduction, the COVID-19 pandemic contributed to weight gain in children in several ways., including increased psychosocial stressors like social isolation and food insecurity, impaired sleep routines, and the promotion of sedentary behaviors(13). In addition to these universally experienced factors, children with a COVID-19 diagnosis may have experienced increased isolation and exposure to traumatic aspects of the pandemic (e.g., having family members pass away from COVID-19), especially early in the pandemic. In addition, for some children, symptoms like fatigue and headaches from a COVID-19 infection may have contributed further to sedentary behaviors. Finally, there is evidence of an increased incidence of Diabetes (both Type 1 and Type 2) with COVID-19 infection because of the pro-inflammatory nature of COVID-19 and its effects on the pancreas(51). While not directly tied to weight gain, it is plausible that these biological factors could have an adverse effect on weight.

Interestingly, among children diagnosed with COVID-19, we found less rapid weight gain in those who were hospitalized compared to those who were not hospitalized. This is consistent with studies in adults demonstrating unintentional weight loss among those hospitalized for COVID-19(52), which may be attributed to inadequate nutritional intake or immobilization during hospitalization. In addition, increased inflammatory cytokines due to severe COVID-19 infection may lead to cachexia and loss of lean body mass(53), as shown in adults. Finally, children hospitalized for COVID-19 comprised only 19% of the total cohort of children diagnosed with COVID-19 in our sample and likely represent more vulnerable populations with more risk for severe disease, whether because of younger age or more medical complexity. Because of their vulnerability, these children may have had closer medical monitoring(54) – especially after hospitalization - that mitigated excessive weight gain.

While the N3C database and our approach to studying the impact of COVID-19 on BMI percentile trajectories in children has several strengths, this study also has limitations to consider. As data are aggregated from many health systems using different CDMs that vary in granularity, some sites had systematic missingness of certain variables. Indeed, our overall sample size decreased from 25,936 children to 11,474 cases due to missing data, especially since we required weight and heights for at least two time-points within a certain timeline compared to diagnosis for both cases and controls to conduct the longitudinal analysis. Furthermore, due to missing data, we used interpolation to fill the gap in the trajectories of %*BMI*_*p*95_ for some patients, which might not represent the actual data. While we adjusted for baseline BMI, the difference in BMI percentiles may still hold clinical significance due to the increased baseline risk of severe outcomes associated with obesity in COVID-19 cases. Given that asymptomatic cases may exhibit distinct weight trajectories compared to symptomatic cases, we opted not to account for asymptomatic cases. This introduces an added layer of complexity to the interpretation of our findings.

We acknowledge that BMI can fluctuate in children over time and that the time window for BMI measurement following diagnosis ranges from one week to six months. This variability could influence the interpretation of the weight trajectories observed, particularly in those with shorter follow-up periods. As such, the generalizability of the findings should be interpreted with caution, especially for participants with only two BMI measurements or shorter time intervals between measurements. Our study did not account for comorbidities associated with sedentary behavior and weight gain, such as diabetes and hypertension, which can influence BMI trajectories and the effect of COVID-19 on weight gain. Future studies should consider these comorbidities to better understand the impact of COVID-19 on weight gain in pediatric populations. These limitations, particularly the lack of adjustment for comorbidities and the focus on U.S.-based data, suggest that caution should be taken when generalizing these results to other populations. Further research is needed to explore the impact of COVID-19 on weight gain in children with different health profiles, particularly in international cohorts and populations not well-represented in the N3C database.

## Conclusion

This study reports on the BMI percentile trajectory of a large nationally representative cohort of children in the United States using the N3C database. Our study uniquely adds to the literature by evaluating how a diagnosis of COVID-19 was associated with BMI percentile trajectories during the pandemic. Children diagnosed with COVID-19 had larger and more persistent increases in BMI percentile over time compared to children without a COVID-19 diagnosis, highlighting the additional impact that a diagnosis of COVID-19 had on weight gain in children beyond the impact of the pandemic generally alone. Future studies should examine the reasons for this association and whether this finding is specific to COVID-19 or could have implications for future health conditions and emergencies.

## Data availability

The data that supports the findings of this study is accessible through the N3C data repository access portal with necessary permissions.

## Acknowledgments

N3C Attribution

The analyses described in this publication were conducted with data or tools accessed through the NCATS N3C Data Enclave https://covid.cd2h.org and N3C Attribution & Publication Policy v 1.2-2020-08-25b supported by NCATS U24 TR002306, Axle Informatics Subcontract: NCATS-P00438-B, and was supported by NIH awards, P20GM103446 and U54-GM104941. This research was possible because of the patients whose information is included within the data and the organizations (https://ncats.nih.gov/n3c/resources/data-contribution/data-transfer-agreement-signatories) and scientists who have contributed to the on-going development of this community resource [https://doi.org/10.1093/jamia/ocaa196].

Disclaimer The N3C Publication committee confirmed that this manuscript msid:1831.353 is in accordance with N3C data use and attribution policies; however, this content is solely the responsibility of the authors and does not necessarily represent the official views of the National Institutes of Health or the N3C program.

IRB

The N3C data transfer to NCATS is performed under a Johns Hopkins University Reliance Protocol # IRB00249128 or individual site agreements with NIH. The N3C Data Enclave is managed under the authority of the NIH; information can be found at https://ncats.nih.gov/n3c/resources.

Individual Acknowledgements for Core Contributors

We gratefully acknowledge the following core contributors to N3C:

Adam B. Wilcox, Adam M. Lee, Alexis Graves, Alfred (Jerrod) Anzalone, Amin Manna, Amit Saha, Amy Olex, Andrea Zhou, Andrew E. Williams, Andrew Southerland, Andrew T. Girvin, Anita Walden, Anjali A. Sharathkumar, Benjamin Amor, Benjamin Bates, Brian Hendricks, Brijesh Patel, Caleb Alexander, Carolyn Bramante, Cavin Ward-Caviness, Charisse Madlock-Brown, Christine Suver, Christopher Chute, Christopher Dillon, Chunlei Wu, Clare Schmitt, Cliff Takemoto, Dan Housman, Davera Gabriel, David A. Eichmann, Diego Mazzotti, Don Brown, Eilis Boudreau, Elaine Hill, Elizabeth Zampino, Emily Carlson Marti, Emily R. Pfaff, Evan French, Farrukh M Koraishy, Federico Mariona, Fred Prior, George Sokos, Greg Martin, Harold Lehmann, Heidi Spratt, Hemalkumar Mehta, Hongfang Liu, Hythem Sidky, J.W. Awori Hayanga, Jami Pincavitch, Jaylyn Clark, Jeremy Richard Harper, Jessica Islam, Jin Ge, Joel Gagnier, Joel H. Saltz, Joel Saltz, Johanna Loomba, John Buse, Jomol Mathew, Joni L. Rutter, Julie A. McMurry, Justin Guinney, Justin Starren, Karen Crowley, Katie Rebecca Bradwell, Kellie M. Walters, Ken Wilkins, Kenneth R. Gersing, Kenrick Dwain Cato, Kimberly Murray, Kristin Kostka, Lavance Northington, Lee Allan Pyles, Leonie Misquitta, Lesley Cottrell, Lili Portilla, Mariam Deacy, Mark M. Bissell, Marshall Clark, Mary Emmett, Mary Morrison Saltz, Matvey B. Palchuk, Melissa A. Haendel, Meredith Adams, Meredith Temple-O’Connor, Michael G. Kurilla, Michele Morris, Nabeel Qureshi, Nasia Safdar, Nicole Garbarini, Noha Sharafeldin, Ofer Sadan, Patricia A. Francis, Penny Wung Burgoon, Peter Robinson, Philip R.O. Payne, Rafael Fuentes, Randeep Jawa, Rebecca Erwin-Cohen, Rena Patel, Richard A. Moffitt, Richard L. Zhu, Rishi Kamaleswaran, Robert Hurley, Robert T. Miller, Saiju Pyarajan, Sam G. Michael, Samuel Bozzette, Sandeep Mallipattu, Satyanarayana Vedula, Scott Chapman, Shawn T. O’Neil, Soko Setoguchi, Stephanie S. Hong, Steve Johnson, Tellen D. Bennett, Tiffany Callahan, Umit Topaloglu, Usman Sheikh, Valery Gordon, Vignesh Subbian, Warren A. Kibbe, Wenndy Hernandez, Will Beasley, Will Cooper, William Hillegass, Xiaohan Tanner Zhang. Details of contributions available at covid.cd2h.org/core-contributors

Data Partners with Released Data

The following institutions whose data is released or pending:

Available: Advocate Health Care Network — UL1TR002389: The Institute for Translational Medicine (ITM) • Aurora Health Care Inc — UL1TR002373: Wisconsin Network For Health Research • Boston University Medical Campus — UL1TR001430: Boston University Clinical and Translational Science Institute • Brown University — U54GM115677: Advance Clinical Translational Research (Advance-CTR) • Carilion Clinic — UL1TR003015: iTHRIV Integrated Translational health Research Institute of Virginia • Case Western Reserve University — UL1TR002548: The Clinical & Translational Science Collaborative of Cleveland (CTSC) • Charleston Area Medical Center — U54GM104942: West Virginia Clinical and Translational Science Institute (WVCTSI) • Children’s Hospital Colorado — UL1TR002535: Colorado Clinical and Translational Sciences Institute • Columbia University Irving Medical Center — UL1TR001873: Irving Institute for Clinical and Translational Research • Dartmouth College — None (Voluntary) Duke University — UL1TR002553: Duke Clinical and Translational Science Institute • George Washington Children’s Research Institute — UL1TR001876: Clinical and Translational Science Institute at Children’s National (CTSA-CN) • George Washington University — UL1TR001876: Clinical and Translational Science Institute at Children’s National (CTSA-CN) • Harvard Medical School — UL1TR002541: Harvard Catalyst • Indiana University School of Medicine — UL1TR002529: Indiana Clinical and Translational Science Institute • Johns Hopkins University — UL1TR003098: Johns Hopkins Institute for Clinical and Translational Research • Louisiana Public Health Institute — None (Voluntary) • Loyola Medicine — Loyola University Medical Center • Loyola University Medical Center — UL1TR002389: The Institute for Translational Medicine (ITM) • Maine Medical Center — U54GM115516: Northern New England Clinical & Translational Research (NNE-CTR) Network • Mary Hitchcock Memorial Hospital & Dartmouth Hitchcock Clinic — None (Voluntary) • Massachusetts General Brigham — UL1TR002541: Harvard Catalyst • Mayo Clinic Rochester — UL1TR002377: Mayo Clinic Center for Clinical and Translational Science (CCaTS) • Medical University of South Carolina — UL1TR001450: South Carolina Clinical & Translational Research Institute (SCTR) • MITRE Corporation — None (Voluntary) • Montefiore Medical Center — UL1TR002556: Institute for Clinical and Translational Research at Einstein and Montefiore • Nemours — U54GM104941: Delaware CTR ACCEL Program • NorthShore University HealthSystem — UL1TR002389: The Institute for Translational Medicine (ITM) • Northwestern University at Chicago — UL1TR001422: Northwestern University Clinical and Translational Science Institute (NUCATS) • OCHIN — INV-018455: Bill and Melinda Gates Foundation grant to Sage Bionetworks • Oregon Health & Science University — UL1TR002369: Oregon Clinical and Translational Research Institute • Penn State Health Milton S. Hershey Medical Center — UL1TR002014: Penn State Clinical and Translational Science Institute • Rush University Medical Center — UL1TR002389: The Institute for Translational Medicine (ITM) • Rutgers, The State University of New Jersey — UL1TR003017: New Jersey Alliance for Clinical and Translational Science • Stony Brook University — U24TR002306 • The Alliance at the University of Puerto Rico, Medical Sciences Campus — U54GM133807: Hispanic Alliance for Clinical and Translational Research (The Alliance) • The Ohio State University — UL1TR002733: Center for Clinical and Translational Science • The State University of New York at Buffalo — UL1TR001412: Clinical and Translational Science Institute • The University of Chicago — UL1TR002389: The Institute for Translational Medicine (ITM) • The University of Iowa — UL1TR002537: Institute for Clinical and Translational Science • The University of Miami Leonard M. Miller School of Medicine — UL1TR002736: University of Miami Clinical and Translational Science Institute • The University of Michigan at Ann Arbor — UL1TR002240: Michigan Institute for Clinical and Health Research • The University of Texas Health Science Center at Houston — UL1TR003167: Center for Clinical and Translational Sciences (CCTS) • The University of Texas Medical Branch at Galveston — UL1TR001439: The Institute for Translational Sciences • The University of Utah — UL1TR002538: Uhealth Center for Clinical and Translational Science • Tufts Medical Center — UL1TR002544: Tufts Clinical and Translational Science Institute • Tulane University — UL1TR003096: Center for Clinical and Translational Science • The Queens Medical Center — None (Voluntary) • University Medical Center New Orleans — U54GM104940: Louisiana Clinical and Translational Science (LA CaTS) Center • University of Alabama at Birmingham — UL1TR003096: Center for Clinical and Translational Science • University of Arkansas for Medical Sciences — UL1TR003107: UAMS Translational Research Institute • University of Cincinnati — UL1TR001425: Center for Clinical and Translational Science and Training • University of Colorado Denver, Anschutz Medical Campus — UL1TR002535: Colorado Clinical and Translational Sciences Institute • University of Illinois at Chicago — UL1TR002003: UIC Center for Clinical and Translational Science • University of Kansas Medical Center — UL1TR002366: Frontiers: University of Kansas Clinical and Translational Science Institute • University of Kentucky — UL1TR001998: UK Center for Clinical and Translational Science • University of Massachusetts Medical School Worcester — UL1TR001453: The UMass Center for Clinical and Translational Science (UMCCTS) • University Medical Center of Southern Nevada — None (voluntary) • University of Minnesota — UL1TR002494: Clinical and Translational Science Institute • University of Mississippi Medical Center — U54GM115428: Mississippi Center for Clinical and Translational Research (CCTR) • University of Nebraska Medical Center — U54GM115458: Great Plains IDeA-Clinical & Translational Research • University of North Carolina at Chapel Hill — UL1TR002489: North Carolina Translational and Clinical Science Institute • University of Oklahoma Health Sciences Center — U54GM104938: Oklahoma Clinical and Translational Science Institute (OCTSI) • University of Pittsburgh — UL1TR001857: The Clinical and Translational Science Institute (CTSI) • University of Pennsylvania — UL1TR001878: Institute for Translational Medicine and Therapeutics • University of Rochester — UL1TR002001: UR Clinical & Translational Science Institute • University of Southern California — UL1TR001855: The Southern California Clinical and Translational Science Institute (SC CTSI) • University of Vermont — U54GM115516: Northern New England Clinical & Translational Research (NNE-CTR) Network • University of Virginia — UL1TR003015: iTHRIV Integrated Translational health Research Institute of Virginia • University of Washington — UL1TR002319: Institute of Translational Health Sciences • University of Wisconsin-Madison — UL1TR002373: UW Institute for Clinical and Translational Research • Vanderbilt University Medical Center — UL1TR002243: Vanderbilt Institute for Clinical and Translational Research • Virginia Commonwealth University — UL1TR002649: C. Kenneth and Dianne Wright Center for Clinical and Translational Research • Wake Forest University Health Sciences — UL1TR001420: Wake Forest Clinical and Translational Science Institute • Washington University in St. Louis — UL1TR002345: Institute of Clinical and Translational Sciences • Weill Medical College of Cornell University — UL1TR002384: Weill Cornell Medicine Clinical and Translational Science Center • West Virginia University — U54GM104942: West Virginia Clinical and Translational Science Institute (WVCTSI) 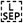 Submitted: Icahn School of Medicine at Mount Sinai — UL1TR001433: ConduITS Institute for Translational Sciences • The University of Texas Health Science Center at Tyler — UL1TR003167: Center for Clinical and Translational Sciences (CCTS) • University of California, Davis — UL1TR001860: UCDavis Health Clinical and Translational Science Center • University of California, Irvine — UL1TR001414: The UC Irvine Institute for Clinical and Translational Science (ICTS) • University of California, Los Angeles — UL1TR001881: UCLA Clinical Translational Science Institute • University of California, San Diego — UL1TR001442: Altman Clinical and Translational Research Institute • University of California, San Francisco — UL1TR001872: UCSF Clinical and Translational Science Institute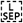 NYU Langone Health Clinical Science Core, Data Resource Core, and PASC Biorepository Core — OTA-21-015A: Post-Acute Sequelae of SARS-CoV-2 Infection Initiative (RECOVER)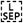 Pending: Arkansas Children’s Hospital — UL1TR003107: UAMS Translational Research Institute • Baylor College of Medicine — None (Voluntary) • Children’s Hospital of Philadelphia — UL1TR001878: Institute for Translational Medicine and Therapeutics • Cincinnati Children’s Hospital Medical Center — UL1TR001425: Center for Clinical and Translational Science and Training • Emory University — UL1TR002378: Georgia Clinical and Translational Science Alliance • HonorHealth — None (Voluntary) • Loyola University Chicago — UL1TR002389: The Institute for Translational Medicine (ITM) • Medical College of Wisconsin — UL1TR001436: Clinical and Translational Science Institute of Southeast Wisconsin • MedStar Health Research Institute — None (Voluntary) • Georgetown University — UL1TR001409: The Georgetown-Howard Universities Center for Clinical and Translational Science (GHUCCTS) • MetroHealth — None (Voluntary) • Montana State University — U54GM115371: American Indian/Alaska Native CTR • NYU Langone Medical Center — UL1TR001445: Langone Health’s Clinical and Translational Science Institute • Ochsner Medical Center — U54GM104940: Louisiana Clinical and Translational Science (LA CaTS) Center • Regenstrief Institute — UL1TR002529: Indiana Clinical and Translational Science Institute • Sanford Research — None (Voluntary) • Stanford University — UL1TR003142: Spectrum: The Stanford Center for Clinical and Translational Research and Education • The Rockefeller University — UL1TR001866: Center for Clinical and Translational Science • The Scripps Research Institute — UL1TR002550: Scripps Research Translational Institute • University of Florida — UL1TR001427: UF Clinical and Translational Science Institute • University of New Mexico Health Sciences Center — UL1TR001449: University of New Mexico Clinical and Translational Science Center • University of Texas Health Science Center at San Antonio — UL1TR002645: Institute for Integration of Medicine and Science • Yale New Haven Hospital — UL1TR001863: Yale Center for Clinical Investigation

## Notes

### Competing Interest Statement

The authors have declared no competing interest.

### Funding Statement

This study was funded by NIH awards, P20GM103446 and U54-GM104941

### Author Declarations

IRB of University of Delaware gave ethical approval for this work.

